# Eruption of COVID-19 like illness in a remote village in Papua (Indonesia)

**DOI:** 10.1101/2020.05.19.20106740

**Authors:** Elco van Burg, Wijnanda van Burg-Verhage

**Author notes:** Corresponding author Emai.

## Abstract

**Background:** The COVID-19 pandemic is creating significant challenges for healthcare infrastructure for countries of all development and resource levels. Low-and-middle resource countries face even larger challenges, as their resources are stretched and often insufficient under normal circumstances. A village in the Papuan highlands of Indonesia; small, isolated, accessed only by small plane or trekking has experienced an outbreak typical of COVID-19.

**Methodology/Principal Findings:** This description was compiled from patient care records by lay healthcare workers in M20 (a pseudonym) during and after an outbreak and from medical doctors responding to online requests for help. We assume that, for reasons given, the outbreak that has been described was COVID-19. The dense social structure of the village resulted in a rapid infection of 90-95% of the population. Physical distancing and isolation measures were used, but probably implemented suboptimal and too late, and their effect on the illness course was unclear. The relatively young population, with a majority of women, probably influenced the impact of the epidemic, resulting in only two deaths so far.

**Conclusions/Significance:** This outbreak pattern of suspected SARS-CoV-2 in a village in the highlands of Papua (Indonesia) presents a unique report of the infection of an entire village population over five weeks. The age distribution, common in Papuan highland villages may have reduced case fatality rate (CFR) in this context and that might be the case in similar remote areas since survival to old age is already very limited and CFR among younger people is lower.

**Author summary:** A village in the Papuan highlands of Indonesia; small, isolated, accessed only by small plane or trekking has experienced an outbreak typical of COVID-19. The outbreak affected 90-95% of approximately 200 residents between 20 February 2020 and 31 March 2020. Lay health workers, consisting of trained local volunteers, without healthcare facilities and limited medication options reached out for help online and managed with what they had available. Their experience is relevant to others in similar settings. This report describes the village, outbreak, timeline, symptoms and treatments, and outcomes.

## Introduction

The COVID-19 pandemic is creating significant challenges for healthcare infrastructure for countries of all development and resource levels. Low-and-middle resource countries face even larger challenges, as their resources are stretched and often insufficient under normal circumstances [1,2]. One such country, Indonesia, is struggling to devise a strategy to manage the COVID-19 pandemic.

The first official SARS-CoV-2 case in Indonesia was detected on 2 March 2020, relatively late, given its close links to the original epicenter, Wuhan. At that time, models had already estimated that there would be more cases [3], and estimated 1-30 directly imported cases from Wuhan [4,5]. In fact, early reports of suspected cases date back to January 2020 [6] Given Indonesia’s health infrastructure challenges, specifically in the remote rural areas of the vast archipelago [7], many predicted that remote spread of SARS-CoV-2 may have severe health effects for these populations.

This case report describes the case of the spread of COVID-19 like illness in one village, M20, in the rural highlands of Papua, Indonesia.

## Methods

This description was compiled from patient care records by lay healthcare workers in M20 during and after an outbreak and from medical doctors responding to online requests for help. We use a pseudonym to disguise the location of the village to protect data of the patients involved, and patient data were analyzed anonymously. The research has been approved by the Research Ethical Review Board of the School of Business and Economics, Vrije Universiteit Amsterdam, The Netherlands. Symptoms of villagers asking for medical help were recorded by lay health workers, so initial, mild symptoms were not systematically recorded. In most cases the exact onset of infection is not clear. A major caveat is that PCR testing for COVID-19 was not possible due to the lack of tests. The team repeatedly contacted the government health services, but PCR testing, or testing using reliable antibody tests, was not possible till the time of this report.

## Results

### Context

M20 is severely isolated at an altitude of 6.700 feet in the central mountain range of Indonesia’s easternmost province, Papua. It is typically served on request by a small 6-8 seat aircraft, or reached by trekking on foot from other villages. The village consists of seven hamlets of 2-6 huts, separated by 5-10 minute walks. Villagers are closely related to inhabitants of other villages in the area, and visit each other often. In Papuan highland villages the men and/or families sleep together in one hut, and children sleep with their mothers or families (sometimes up to 30 people in one hut with visitors). The closest village geographically is about a 1.5 hour hike down the mountain, with social interaction multiple times a week. The actual population varies with these interactions ranging between 150-200 people. Approximately half of the population is under 12 years of age. There are 4-6 matriarchs and the rest are teenagers, young adults and adults in their 30s-50s. M20 gender distribution is estimated at 60% women, and 40% men, due to higher life expectancy of women (66.8 compared to 63.0 for men) [8] and men spending the majority of their time in towns.

The closest government health center is about three hours hike away, but the trained health worker is typically absent, as is common in this region [9]. Lay health workers do daily clinics with basic medicine. They report that common health problems include ear infections and pneumonia as the main sicknesses. The village altitude is too high for endemic malaria, but some malaria is imported from towns and villages at lower elevation. Most men smoke, and most people live in huts with central fire pits, which are used throughout the day for cooking and during the evening and night for heating [10] TB and HIV have not been diagnosed by the lay health workers in M20.

### Epidemiologic timeline

The index patient in this outbreak reported for care on 20 February, with symptoms of what, in hindsight, is suspected COVID-19. This patient might have been the first to bring it to the village, as he had traveled to a neighboring area reporting similar symptoms. Alternatively, airplane visitors with links to Jakarta or Jayapura are suspected to have transmitted the virus (in both cities COVID-19 symptoms have been reported in February). Two weeks after this index case, the medical workload for the local healthcare workers rose steeply (see Figure 1).

**Figure 1.**
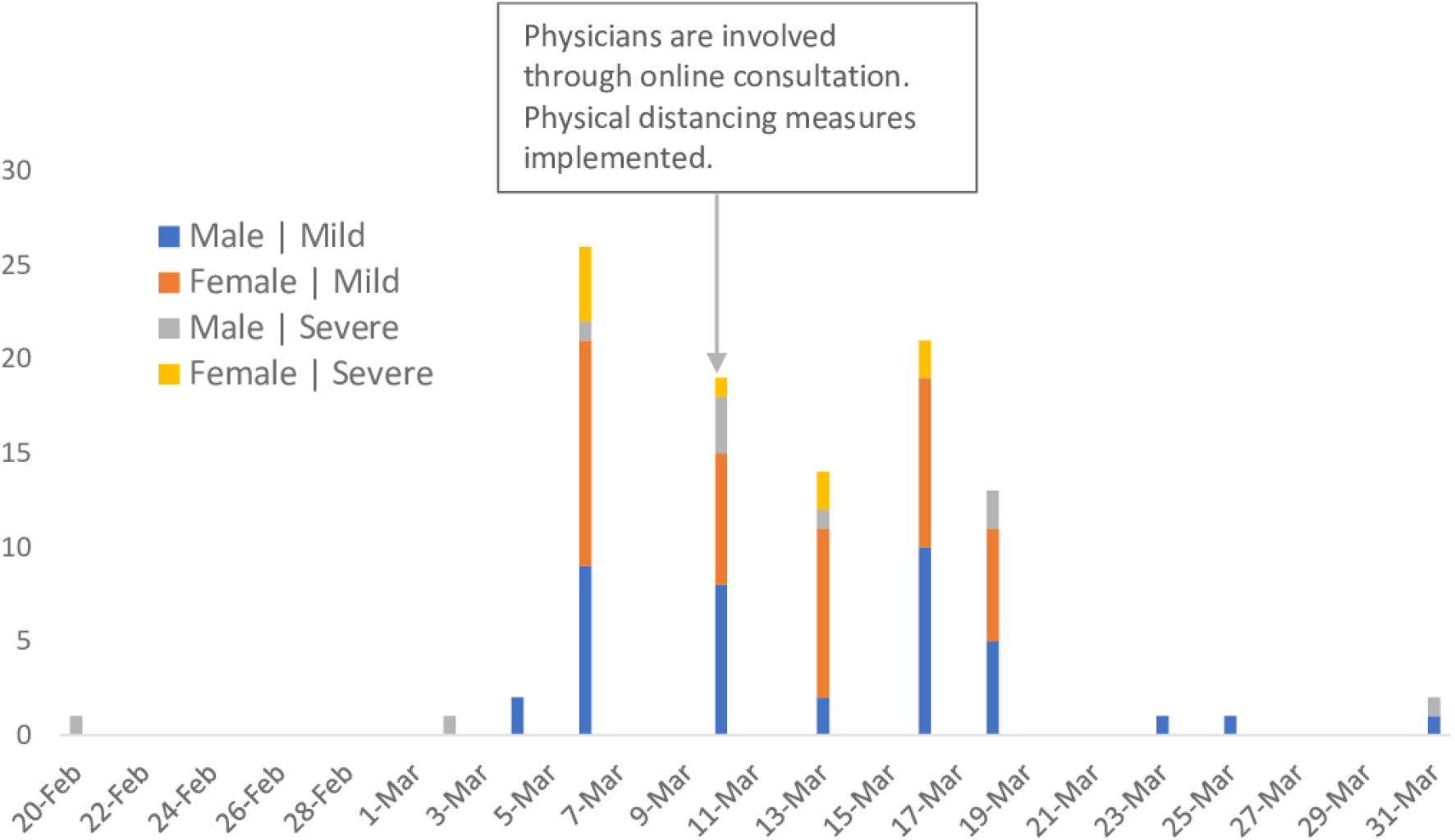
Unique Patients with Symptoms Observed over Time.

The patients reported symptoms typical of COVID-19. Clinic records showed the following symptoms as summarized in Table 1. Table 2 presents the characteristics of the patients.

**Table 1.** Suspected COVID-19 Patients Treated in M20.

| Age | Mild to moderate symptoms Male | Mild to moderate symptoms Female | Severe symptoms Male | Severe symptoms Female |
| --- | --- | --- | --- | --- |
| 0-5 | 7 6.9% | 5 5.0% | 2 2.0% | 2 2.0% |
| 6-10 | 7 6.9% | 15 14.9% | 1 1.0% | 1 1.0% |
| 11-15 | 2 2.0% | 2 2.0% | 0 0.0% | 1 1.0% |
| 16-20 | 8 7.9% | 1 1.0% | 0 0.0% | 0 0.0% |
| 21-25 | 2 2.0% | 2 2.0% | 1 1.0% | 0 0.0% |
| 26-30 | 4 4.0% | 3 3.0% | 1 1.0% | 3 3.0% |
| 31-35 | 3 3.0% | 6 5.9% | 1 1.0% | 0 0.0% |
| 36-40 | 3 3.0% | 5 5.0% | 1 1.0% | 0 0.0% |
| 41-45 | 1 1.0% | 3 3.0% | 0 0.0% | 0 0.0% |
| 46-50 | 2 2.0% | 1 1.0% | 3 3.0% | 1 1.0% |
| 50+ | 0 0.0% | 0 0.0% | 0 0.0% | 1 1.0% |
| Mean=21.7 |  |  |  |  |
| Total | 39 38.6% | 43 42.6% | 10 10.1% | 9 8.9% |

**Table 2.** Symptoms of Patients with Suspected COVID-19.

| **Symptom** | **Approximate Percentage of Patients Reporting** |
| --- | --- |
| Fever (either mild 37-38.5C or High < 39.5) | 80% |
| Sore or dry throat | 90-95% Adults. 60-70% children |
| Coughing (usually at night) | 80% |
| Fatigue | 90% |
| Lethargy (most of the children) | Many |
| Headache late in the illness | 60% |
| Muscle and joint pain | 10-20% |
| Shortness of Breath | 70% |
| Diarrhea | 20-30% |
| Vomiting | 10% |

The illness started with several days of sore throats, followed by stomach complaints (diarrhea and vomiting), and next fever within 24 hours of stomach complaints. Fever and fatigue were constant, lasting 3-5 days. In severe cases, fevers above 40°C often accompanied shortness of breath and chest pain. Severe symptoms generally occurred after day five of illness. Two villagers died on 9 March. Both were male, over 40, and had apparent underlying chronic illness (most likely kidney problems). They had 48 hours of extreme shortness of breath.

Since testing was not available, the following differential diagnoses were considered based on clinical characteristics. 1) COVID-19, symptoms and rapid spread of the illness match best with known symptoms of COVID-19. 2) Influenza: symptoms were consistent. In influenza spread is typically less rapid [11,12] 3) RSV (or other virus): RSV usually hits babies the hardest. which was not true in this illness. 4) Bacterial pneumonia, which is common, especially in children [13,14] expect low human to human transmission, also antibiotics did not affect speed of recovery. 5) Pertussis: epidemic pertussis had affected this village in 2019 with a subsequent vaccine campaign. The characteristic cough in this illness was less intense than typical pertussis.

The healthcare team treated symptoms with paracetamol, up to four times a day. Temperatures were checked daily. Patients considered ‘very sick’, with climbing temperatures, diminished or crackling lung sounds, or ear infections (which are very common in Papua’s highland population anyway) were given empiric Amoxicillin treatment (49% of 101 patients) to prevent/treat secondary pneumonia. Those with fevers over 40°C received a different antibiotic (Amoxicillin/Clavulanic Acid (2% of the patients) or Azithromycin (3% of the patients)). The relationship of antibiotics to recovery is not clear.

Because of the increased medical workload, and two deaths in the village, workers reached out online for medical support to physicians specialized in tropical medicine on 13 March. Suspecting a virus, maybe SARS-CoV-2, consulting physicians advised them to implement physical distancing measures. Implementation was suboptimal with exposure already significant in the community.

Upon learning that chloroquine might help stop coronavirus from attaching to the receptors [15], people in the ‘very sick’ group and high risk people (ages 40 + and those that were often sick), were given 150mg chloroquine (250mg pill) twice a day for seven days (14% of the patients). Two older ladies (2% of the cases) were ‘very sick’ and were also given Azithromycin (500mg initially, 250mg the next four days) based on reports that it might help. Except the two who died, all patients are recovering or have recovered.

As Figure 1 shows, the entire epidemic curve covers five weeks. The healthcare team treated 101 patients (half the village). Informal questioning in the community revealed that only about 10 villagers denied any symptoms yielding a presumptive infection rate of 90-95% of all residents within that 4-week period. Table 1 shows that 5 of the 12 patients over age 40 were very sick, and two of them died. This yields an overall 41% severe and 17% mortality rate among patients over age 40 and roughly corresponds with other reports [16–18] This very young age for elderly members of the community reflects the inherent difficulty of survival in this region. Approximately 50% of villagers experienced mild or no symptoms.

## Discussion and conclusion

This outbreak pattern of suspected SARS-CoV-2 in a village in the highlands of Papua (Indonesia) presents a unique report of the infection of an entire village population over five weeks. We assume that, for the reasons given, the outbreak that has been described was COVID-19. The dense social structure of the village resulted in a rapid infection of 90-95% of the population. Physical distancing and isolation measures were used, but probably implemented suboptimal and too late, and their effect on the illness course was unclear.

The M20 population is uniquely young with a mean patient age of 21.7. This might partially explain the unexpectedly low overall death rate of 1% in the context of minimal health facilities and no mitigating measures. Two women, over age 50, got severely ill and were critically affected by the disease but survived. This age distribution, common in Papuan highland villages may reduce case fatality rate (CFR) in similar remote areas since survival to old age is already very limited and CFR among younger people is lower. However, the lives and wisdom of these surviving elders needs to be carefully protected and guarded for tribal, cultural, and social survival. Treatment with chloroquine phosphate and azithromycin were used, but caution and further research are needed [19].

A high proportion of women in the demographics may explain that in this village women were affected more often by the disease than men, but in the older age groups, men tended to be very sick more often and had higher mortality than women in their age cohorts, similar to general epidemiologic data of COVID-19.

## Data Availability

Data are included in the manuscript.

